# Dietary Intake During breast cancer treatment and Its Association with Metabolic and Psychological Outcomes in Women with Breast Cancer: A Prospective Cohort Study Protocol

**DOI:** 10.64898/2026.07.20.26358546

**Authors:** Omid Asbaghi, Mohammad Esmaeil Akbari, Hamid Reza Mirzaei, Bahareh Nikooyeh, Seyed Hossein Davoodi

**Author notes:** **Corresponding Author:** Seyed Hossein Davoodi, National Nutrition and Food Technology Research Institute, Shahid Beheshti University of Medical Sciences (SBUMS), Tehran, Iran.

## Abstract

**Background:** Active systemic therapy, including chemotherapy alone or combined with HER2-directed targeted therapy, is a cornerstone of breast cancer management, yet it frequently precipitates adverse cardiometabolic, anthropometric, and psychological disturbances. Although dietary intake during active treatment may modulate these complications, rigorous prospective cohort evidence utilizing repeated dietary assessments and standardized clinical endpoints remains limited.

**Objective:** The primary objective of this prospective cohort study is to investigate the longitudinal association between dietary intake during active cancer treatment and changes in the visceral adiposity index (VAI), serving as the primary cardiometabolic endpoint, among women with breast cancer. Secondary objectives are to examine associations between dietary exposures and individual cardiometabolic markers, body composition, psychological distress, and health-related quality of life.

**Methods:** A prospective cohort of 100 women with histologically confirmed primary breast cancer is recruited at the initiation of systemic therapy and followed prospectively for six months in Tehran, Iran. The cohort encompasses patients undergoing standard chemotherapy regimens (spanning 4.5 to 6 months), including those receiving sequential HER2-directed targeted therapy. Habitual dietary intake during active treatment is assessed using nine 24-hour dietary records systematically distributed across the early, middle, and late treatment phases (incorporating weekdays and weekend days). Clinical, anthropometric, biochemical, and psychological outcomes are evaluated at baseline (pre-treatment) and at the 6-month follow-up. Primary multivariable linear regression models will specify the 6-month VAI as the dependent variable, adjusting for its baseline level, age, baseline body mass index, tumor stage, active treatment regimen, follow-up physical activity, and total energy intake. Secondary analyses will evaluate key secondary cardiometabolic parameters, psychological outcomes, and exploratory end-of-study serum 25-hydroxyvitamin D concentrations. Regression results will be reported as unstandardized coefficients (*B*) with 95% confidence intervals and standardized coefficients (*β*).

**Conclusion:** This prospective protocol outlines a standardized methodology to determine whether dietary intake during active systemic therapy tracks with adverse cardiometabolic trajectories and psychological burden, generating crucial evidence to inform tailored nutritional interventions during active breast cancer treatment.

## Introduction

Breast cancer is the most commonly diagnosed cancer among women worldwide and remains a leading cause of cancer-related morbidity and mortality ^1^. Advances in early detection and systemic treatments have substantially improved survival rates; however, cytotoxic chemotherapy, alone or combined with targeted agents, remains a cornerstone of therapy for many patients ^2^. Despite its therapeutic benefits, systemic treatment is frequently accompanied by adverse effects, including metabolic disturbances, alterations in body composition, dyslipidemia, cardiovascular complications, hematological changes, and psychological distress ^3^, all of which may negatively affect patients’ quality of life and long-term prognosis.

Accumulating evidence demonstrates that systemic cancer therapy can induce unfavorable cardiometabolic alterations, such as weight gain, increased visceral adiposity, insulin resistance, and lipid abnormalities. Concurrently, patients frequently experience hematological toxicities, hormonal shifts, fatigue, anxiety, depression, and marked declines in health-related quality of life. These treatment-induced perturbations not only undermine immediate therapy tolerance and psychological well-being during active treatment, but also exacerbate long-term cardiometabolic risk and chronic disease vulnerability in this population ^4–14^.

Nutrition is increasingly recognized as a critical modifiable factor that may influence treatment tolerance, metabolic resilience, and clinical trajectories in oncology. During active cancer treatment, changes in appetite, taste perception, and gastrointestinal function frequently lead to inadequate or imbalanced nutrient intake, potentially modulating systemic inflammation, metabolic responses, and psychological well-being. However, existing studies are largely constrained by single-point dietary assessments and a narrow scope of clinical endpoints. Prospective evidence evaluating repeated dietary intake throughout the entire active treatment period alongside comprehensive metabolic, hematological, and psychological outcomes remains scarce, particularly in Middle Eastern populations.

Therefore, this prospective cohort study is designed to investigate the association between dietary intake during active systemic treatment (chemotherapy with or without targeted therapy) and longitudinal changes in cardiometabolic risk profiles, with the Visceral Adiposity Index (VAI) serving as the primary cardiometabolic endpoint, in women with breast cancer. Secondary analyses will examine associations between dietary exposures and key secondary outcomes, including lipid profile, body composition parameters, glycemic and liver markers, as well as psychological distress and health-related quality of life. By combining repeated 24-hour dietary records with structured longitudinal clinical and patient-reported assessments over a 6-month follow-up, this study aims to clarify the role of diet as a modifiable determinant of cardiometabolic and psychological outcomes during cancer therapy.

## Literature Review

### Chemotherapy-related metabolic and psychological changes in breast cancer

Systemic cancer therapy, encompassing chemotherapy and targeted agents, remains a central component of breast cancer management in both adjuvant and neoadjuvant settings. A growing body of evidence documents the substantial metabolic and psychological burden associated with active systemic treatment, providing the empirical basis for investigating dietary intake as a modifiable determinant of these outcomes ^13^.

A consistent finding across prospective studies is the emergence of metabolic syndrome (MetS) and its individual components during chemotherapy. (Neo)adjuvant regimens have been shown to significantly increase body weight, fat mass, atherogenic lipids, and glucose-metabolism dysregulation in women free of MetS at baseline ^10^, with marked deterioration in fasting insulin, HOMA-IR, and HbA1c reported across cohorts ^5^, particularly in premenopausal patients. Larger cohort studies have documented persistent increases in waist circumference, dyslipidemia ^6^, and a heightened incidence of MetS and type 2 diabetes following treatment, especially in triple-negative subtypes ^12^. rucially, these metabolic shifts are multifactorial, driven by an interplay between cytotoxic agent toxicities, supportive medications (e.g., corticosteroids), and concurrent lifestyle disruptions, notably dietary shifts and physical inactivity.

Evidence regarding insulin dynamics and adipokine responses during active cancer treatment is heterogeneous, largely reflecting differences in therapeutic regimens, menopausal status, and cancer stage. Substantial increases in circulating insulin, resistin, and HOMA-IR have been reported ^9^, alongside strong correlations between leptin and HOMA-IR during taxane-based regimens ^7^. In contrast, attenuated post-load insulin responses have been observed in advanced/stage IV cohorts ^4^. a divergence likely driven by advanced disease-related catabolism rather than cytotoxic effects per se. These inconsistencies underscore the critical need for prospective studies in homogeneous, non-metastatic cohorts that rigorously account for specific regimen types, treatment duration, and baseline metabolic status.

Beyond metabolic disturbances, systemic cancer therapy imposes a profound psychological burden. Clinically meaningful levels of anxiety, depressive symptoms, and impaired health-related quality of life are documented across active treatment phases, frequently compounded by clinical factors such as disease stage, age, and treatment intensity ^14^. Severe fatigue, cognitive complaints, and sleep fragmentation further exacerbate emotional distress and interact dynamically with nutritional intake, establishing a bidirectional relationship between dietary behaviors, metabolic status, and psychological well-being throughout active therapy.

The consistency of treatment-associated weight gain, dyslipidemia, and insulin resistance provides a strong rationale for examining dietary intake as a primary modifiable exposure. Evidence from human metabolic studies demonstrates that macronutrient quality and overall diet composition directly influence visceral adiposity and ectopic fat accumulation independently of total energy intake and absolute weight change ^11^. This indicates that dietary quality during active cancer treatment may directly modulate surrogate visceral adiposity markers (such as the Visceral Adiposity Index) and cardiometabolic risk trajectories, though prospective empirical evidence in breast cancer cohorts undergoing active treatment remains scarce.In summary, chemotherapy contributes to metabolic and psychological deterioration in women with breast cancer, but most existing studies are limited by small samples, heterogeneous regimens, and the absence of detailed dietary assessment. Whether dietary intake during chemotherapy modifies these treatment-related changes remains an open question that the present study is designed to address.

In summary, systemic therapy contributes to marked metabolic and psychological deterioration in women with breast cancer; however, existing literature is predominantly constrained by small sample sizes, cross-sectional designs, and an absence of detailed, longitudinal dietary tracking. Whether sustained dietary quality and specific macronutrient distributions during active treatment mitigate these treatment-induced adverse trajectories remains a critical question that the present prospective cohort study is designed to address.

### Nutrition during chemotherapy in breast cancer

Malnutrition, muscle mass depletion, and metabolic alterations are highly prevalent during cancer treatment and adversely influence treatment tolerance, toxicities, and clinical outcomes 15. International oncology nutrition guidelines, including those from the European Society for Clinical Nutrition and Metabolism (ESPEN), strongly recommend systematic malnutrition screening and stepwise nutritional interventions ranging from individualized dietary counseling to targeted supplementation—combined with physical activity as an integral standard of oncologic care ^15^.

Prospective data in women with breast cancer indicate that active systemic therapy frequently leads to a deterioration in overall diet quality. In a longitudinal cohort of 55 patients, treatment was accompanied by declining intake of fruits, vegetables, and legumes, alongside marked reductions in essential micronutrient intake and a high prevalence of nutrient inadequacies, despite concurrent increases in body weight, BMI, and central adiposity ^16^. These observations demonstrate that cancer therapy can paradoxically impair dietary nutrient density while simultaneously promoting adiposity gain. Qualitative and observational findings similarly indicate that patients frequently enter treatment with suboptimal nutritional patterns and struggle to maintain balanced nutrient quality amidst escalating fatigue and reduced physical activity ^17^.

Treatment-induced chemosensory alterations represent a primary physiological mechanism linking active therapy to nutritional deterioration. Taste and smell alterations have been shown to significantly compromise nutritional status, with higher chemosensory symptom severity independently predicting poorer Mini Nutritional Assessment scores in oncology cohorts ^18^. When combined with treatment-related nausea, gastrointestinal discomfort, and oral mucositis, these sensory alterations drive selective food aversions and reduced dietary diversity ^16^ ^17^, substantially compounding the nutritional challenges during active treatment.

Interventional evidence demonstrates that structured nutritional support during active cancer therapy is both feasible and clinically beneficial. A systematic review of individualized dietary counseling during chemotherapy reported significant improvements in nutritional status, body composition trajectories, cancer-related fatigue, global quality of life, and treatment adherence in intervention groups ^19^. Furthermore, systematic evaluations of oral nutritional supplements have shown that formulations enriched with high-quality whey protein, branched-chain amino acids, or omega-3 polyunsaturated fatty acids assist in preserving lean mass and reducing specific treatment toxicities, such as severe oral mucositis ^20^.

Beyond absolute energy balance, overall dietary pattern quality and macronutrient distribution are critical determinants of patient outcomes. High adherence to nutrient-dense dietary patterns, such as the Mediterranean diet, is associated with favorable cardiometabolic profiles and improved survival trajectories in oncology populations ^21^. Among breast cancer patients, higher post-diagnosis diet quality has been directly linked to superior physical and mental health-related quality of life, independent of key clinical confounders ^22^. Furthermore, oncology guidelines emphasize maintaining adequate protein intake (typically 1.0 to 1.5 g/kg/day) to prevent skeletal muscle wasting and support metabolic resilience throughout active cytotoxic treatment ^15^.

Taken together, current evidence indicates that systemic treatment for breast cancer is frequently accompanied by declines in diet quality, inadequate essential nutrient density, weight gain, and treatment-related sensory toxicities that impair overall nutritional status. While higher-quality dietary patterns and adequate macronutrient intake are linked to improved clinical trajectories, the majority of existing prospective studies rely on single, cross-sectional dietary evaluations and track limited clinical endpoints. The present study addresses this critical gap through rigorous, repeated dietary assessments (nine 24-hour dietary records across treatment phases) and comprehensive longitudinal evaluations of cardiometabolic and psychological outcomes throughout active treatment.

## Methods

### Study design and participants

“This prospective cohort study is conducted in Tehran, Iran, among 100 women with histologically confirmed breast cancer initiating active systemic therapy. All participants are followed prospectively for a fixed period of 6 months from treatment initiation. The active treatment course spans 4.5 to 6 months of chemotherapy; for HER2-positive participants completing a 4.5-month chemotherapy course, the remaining 1.5 months within this observation window comprise HER2-directed targeted therapy (e.g., trastuzumab), ensuring a uniform 6-month follow-up. The primary objective is to evaluate the association between longitudinal dietary intake during treatment and changes in cardiometabolic parameters (primarily the Visceral Adiposity Index [VAI]), body composition, and psychological outcomes. Dietary intake is assessed longitudinally using nine 24-hour dietary recalls across treatment phases. Clinical, anthropometric, psychological, and laboratory measures are evaluated at two standardized time points: baseline (prior to systemic therapy) and at the 6-month follow-up (marking active treatment completion). Predefined outcome hierarchies are systematically applied to guide statistical inferences.”

**The primary outcome** is visceral adiposity index (VAI).

**The key secondary outcomes** are fat mass (FM), homeostatic model assessment of insulin resistance (HOMA-IR), and aspartate aminotransferase (AST).

**Other secondary outcomes** include waist circumference (WC), triglycerides (TG), low-density lipoprotein cholesterol (LDL-C), high-density lipoprotein cholesterol (HDL-C), systolic blood pressure (SBP), diastolic blood pressure (DBP), fasting blood glucose (FBG), glycated hemoglobin (HbA1c), fasting insulin, alanine aminotransferase (ALT), and psychological outcomes.

### Sample Size Calculation

The sample size was determined a priori for the prospective cohort study, designed to evaluate associations between treatment-period dietary intake and cardiometabolic outcomes using multivariable linear regression. Sample size estimation was performed using G*Power software (version 3.1.9.2) based on an F-test for multiple linear regression (*R*^2^ deviation from zero), assuming a medium effect size (*f*^2^ 0.20), a two-sided significance level (*α*) of 0.05, 80% statistical power, and seven independent predictors. Under these parameters, a minimum sample size of 80 participants was required. To account for an anticipated 20% attrition rate over the 6-month active treatment course, target recruitment was established at 100 participants.

### Eligibility Criteria

Participants were included if they met all of the following baseline criteria: (i) histologically confirmed primary breast cancer; (ii) scheduled to initiate active systemic chemotherapy; (iii) aged between 18 and 75 years; and (iv) willingness and cognitive ability to complete dietary records and study questionnaires. Baseline exclusion criteria comprised: (i) pregnancy or lactation; (ii) adherence to a highly restrictive diet (self-reported total energy intake <500 kcal/day); (iii) pre-existing chronic conditions substantially altering metabolic or liver parameters (e.g., pre-existing diabetes mellitus, chronic renal failure, active liver cirrhosis, or major gastrointestinal malabsorptive disorders); (iv) regular use of medications, high-dose dietary supplements, or herbal products known to fundamentally alter glucose or lipid metabolism; and (v) history of substance abuse. Participants who subsequently experienced major protocol deviations, withdrew consent, failed to complete the 6-month follow-up assessments, or provided fewer than seven valid dietary records were documented as study withdrawals and excluded from final complete-case analyses.

### Demographic and clinicopathological characteristics

Baseline demographic and clinicopathological data are collected prior to the first chemotherapy cycle via face-to-face structured interviews and medical record reviews. Demographic characteristics include age, menopausal status, educational attainment, marital status, employment status, smoking history, parity, and family history of cancer. Clinicopathological parameters extracted from medical charts comprise tumor stage, histological grade, tumor size, lymph-node involvement, metastatic status, and hormone receptor status, including estrogen receptor (ER), progesterone receptor (PR), and human epidermal growth factor receptor 2 (HER2). Treatment-related variables include the clinical setting (adjuvant vs. neoadjuvant), specific chemotherapy regimen, and the receipt of concurrent or sequential HER2-directed targeted therapy. The administered systemic regimens encompass: (i) AC (doxorubicin plus cyclophosphamide; four cycles every three weeks), (ii) T (docetaxel; four cycles every three weeks, sequential to AC), (iii) TC (docetaxel plus cyclophosphamide; four to six cycles), and (iv) HER2-directed targeted regimens incorporating trastuzumab with or without pertuzumab, administered according to institutional oncologic guidelines.

### Dietary Intake Assessment

Dietary intake is longitudinally assessed using repeated 24-hour dietary records collected throughout the active treatment course. Each participant completes nine records in total, systematically distributed across three distinct treatment intervals: early, middle, and late chemotherapy phases (three records per phase, incorporating two weekdays and one weekend day per phase). To mitigate the acute distortion of infusion toxicities and capture habitual dietary patterns, records are strictly collected on non-chemotherapy days, excluding the day of drug administration as well as the immediately preceding and subsequent days. Participants receive standardized individual training using visual aids for portion size estimation, household measures, and commercial brand reporting. All records are reviewed face-to-face or verified by telephone interviews by trained clinical nutritionists. Daily nutrient intakes are quantified using Nutritionist IV software adapted for Iranian food items. The arithmetic mean of all valid records represents overall dietary exposure during treatment. A minimum of seven valid records out of nine is required for inclusion in the per-protocol dietary analyses.

### Definition of Dietary Exposures

Prespecified dietary exposures encompass total energy, macronutrients (protein, fat, carbohydrates, and dietary fiber), selected micronutrients (vitamins C, D, E, folate; calcium; iron; zinc), major food groups (fruits, vegetables, whole grains, dairy, red/processed meats), and validated dietary quality scores. For each secondary hypothesis testing, exposures are designated a priori based on biological plausibility and pre-specified in the statistical analysis plan. Absolute intakes of micronutrients and macronutrients are energy-adjusted using the nutrient residual method, or expressed as percentage of total energy intake (%E) for macronutrient distribution models, to isolate dietary composition effects from total energy intake.

### Physical Activity Assessment

Physical activity is assessed at baseline and at the 6-month follow-up using the validated Persian short form of the International Physical Activity Questionnaire (IPAQ-SF), expressed as continuous metabolic equivalent task (MET)-minutes per week. Although the IPAQ-SF relies on self-report, it offers an established, feasible metric in oncology populations. Total physical activity at the 6-month follow-up serves as the primary concurrent lifestyle covariate in multivariable models. Baseline physical activity is retained for sensitivity analyses to verify whether adjusting for pre-treatment activity alters observed longitudinal associations.

### Anthropometric and Body-Composition Assessment

Anthropometric and body-composition parameters are measured at baseline (prior to systemic therapy) and at the 6-month follow-up by trained researchers using standardized protocols. Body weight is measured to the nearest 0.1 kg with participants barefoot and wearing light clothing, using a calibrated digital scale, and standing height is recorded to the nearest 0.1 cm using a wall-mounted stadiometer. Body mass index (BMI) is calculated as weight in kilograms divided by height in meters squared (*kg/m*^2^). Waist circumference (WC) is measured at the midpoint between the lower costal margin and the iliac crest using an inelastic measuring tape in accordance with WHO guidelines. Whole-body fat mass (FM, in kg and percentage) is assessed using multi-frequency bioelectrical impedance analysis (BIA) under standardized fasting and hydration conditions. Blood pressure and resting heart rate are measured in duplicate after a 5-minute seated rest using a clinically validated automated sphygmomanometer, with the average of two concordant readings recorded.

### Blood Sampling and Laboratory Measurements

Venous blood samples are drawn after an overnight 10–12 hour fast at baseline (prior to systemic therapy) and at the 6-month follow-up. Serum and plasma fractions are separated via centrifugation within 45 minutes of collection and stored in aliquots at −80°C until batched analysis in a single accredited central laboratory. Direct automated biochemical assays determine fasting blood glucose (FBG), glycated hemoglobin (HbA1c), fasting insulin, lipid profiles (triglycerides [TG], total cholesterol, low-density lipoprotein cholesterol [LDL-C], and high-density lipoprotein cholesterol [HDL-C]), as well as liver enzymes (aspartate aminotransferase [AST] and alanine aminotransferase [ALT]). Serum 25-hydroxyvitamin D [25(OH)D] is quantified at the 6-month follow-up using a standardized enzyme-linked immunosorbent assay (ELISA) and evaluated as an exploratory concurrent biomarker, with explicit recognition of its cross-sectional limitation at study completion.

### Psychological Outcomes and Quality-of-Life Assessment

Psychological distress and health-related quality of life are evaluated longitudinally at baseline and at the 6-month follow-up. Depressive symptoms, anxiety, and perceived stress are assessed using the validated Persian version of the 21-item Depression, Anxiety and Stress Scale (DASS-21). Health-related quality of life is evaluated using the European Organisation for Research and Treatment of Cancer core questionnaire (EORTC QLQ-C30) alongside the breast cancer– specific module (QLQ-BR23). Questionnaires are administered through in-person structured interviews by trained staff, with standardized telephone follow-up employed when necessary to minimize missing items.

### Outcome Domains

Study endpoints are systematically classified into two predefined domains with a hierarchical structure designed to mitigate multiplicity and control Type I error inflation.

- Domain 1: Cardiometabolic and Biochemical Outcomes

`o` *Primary Outcome:* The primary outcome is the Visceral Adiposity Index (VAI), a validated sex-specific surrogate index of visceral adipose tissue dysfunction, calculated from anthropometric and lipid parameters
`o` *Secondary Outcomes:* Waist circumference (WC), Whole-body fat mass (FM), triglycerides (TG), low-density lipoprotein cholesterol (LDL-C), high-density lipoprotein cholesterol (HDL-C), systolic blood pressure (SBP), diastolic blood pressure (DBP), fasting blood glucose (FBG), glycated hemoglobin (HbA1c), fasting serum insulin, and alanine aminotransferase (ALT), homeostatic model assessment of insulin resistance (HOMA-IR), and aspartate aminotransferase (AST).
- Domain 2: Psychological and Quality-of-Life Outcomes

`o` Secondary endpoints in this domain encompass emotional distress (depressive symptoms, anxiety, and perceived stress evaluated via DASS-21) and multi-dimensional health-related quality of life (global health status, functional domains, and symptom burden evaluated via EORTC QLQ-C30 and QLQ-BR23).

### Calculation of Derived Indices

#### Adiposity, glycemic and lipid-related indices will be calculated using the following formulas

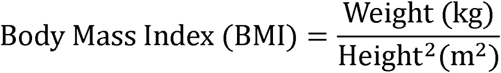

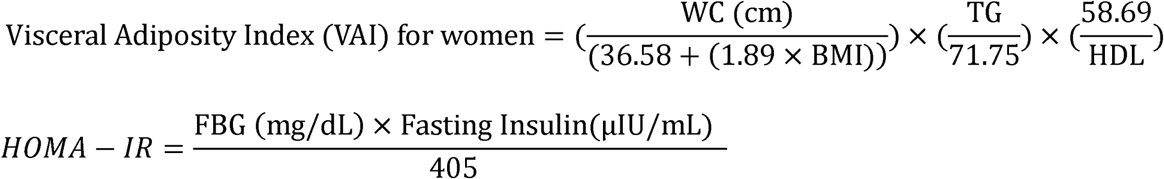

Where WC is waist circumference in cm, BMI is in kg/m, and fasting TG and HDL-C are in mg/dL.

### Serum Vitamin D Analysis

This prospective cohort study is conducted in Tehran, Iran, among 100 women with histologically confirmed breast cancer initiating active systemic therapy. All participants are followed prospectively for a fixed period of 6 months from treatment initiation. The active treatment course spans 4.5 to 6 months of chemotherapy; for HER2-positive participants completing a 4.5-month chemotherapy course, the remaining 1.5 months within this observation window comprise HER2-directed targeted therapy (e.g., trastuzumab), ensuring a uniform 6-month follow-up. The primary objective is to evaluate the association between longitudinal dietary intake during treatment and changes in cardiometabolic parameters (primarily the Visceral Adiposity Index [VAI]), body composition, and psychological outcomes. Dietary intake is assessed longitudinally using nine 24-hour dietary recalls across treatment phases. Clinical, anthropometric, psychological, and laboratory measures are evaluated at two standardized time points: baseline (prior to systemic therapy) and at the 6-month follow-up (marking active treatment completion). Predefined outcome hierarchies are systematically applied to guide statistical inferences.

### Data Management and Quality Control

Data will be collected using standardized forms and entered into a secure electronic database by trained researchers. Each participant will be assigned a unique identification code, and personally identifiable information will be stored separately from the analytical database. Range checks, logical consistency assessments, and comparisons with the original data-collection forms will be performed before statistical analysis.

Patterns and proportions of missing data will be examined before conducting statistical tests. Participants with fewer than seven valid dietary records will be considered to have missing dietary exposure data. When the proportion of missing covariate or outcome data is low, complete-case analysis will be used. If missingness exceeds approximately 5% and appears compatible with the missing-at-random assumption, multiple imputation by chained equations will be considered, with complete-case analysis retained as a sensitivity analysis.

### Statistical Analysis

#### General Statistical Approach

All statistical analyses will be performed using SPSS software (version 25.0; IBM Corp., Armonk, NY, USA). Continuous variables will be summarized as mean ± standard deviation (SD) for approximately normally distributed data, or as median with interquartile range (IQR) for skewed distributions. Categorical variables will be expressed as absolute frequencies and percentages. The distribution and normality of continuous variables will be assessed using the Shapiro–Wilk test alongside visual inspection of histograms and normal Q–Q plots; variables exhibiting marked skewness will undergo logarithmic transformation prior to parametric procedures. Baseline characteristics and unadjusted bivariate associations will be compared using independent-samples *t*-tests or Mann–Whitney *U* tests for continuous variables, and Pearson’s chi-square or Fisher’s exact tests for categorical variables. Longitudinal within-group changes from baseline to the 6-month follow-up will be evaluated using paired-samples *t*-tests or Wilcoxon signed-rank tests, as appropriate. All statistical tests will be two-tailed, and a *P*-value < 0.05 will be considered statistically significant.

##### Note on dietary exposure operationalization

To capture habitual dietary intake throughout active systemic therapy and minimize day-to-day within-person variation, dietary exposures are calculated as the arithmetic mean across all valid 24-hour dietary records. Macronutrient intakes are expressed as percentage of total energy intake (%E), whereas absolute micronutrient and food group intakes are energy-adjusted using the nutrient residual method prior to downstream multivariable modeling.

#### Analysis 1: Changes in outcomes during active cancer treatment

Longitudinal changes in anthropometric, cardiometabolic, and psychological parameters will be calculated as the 6-month follow-up value minus the baseline pre-treatment value (Δ follow-up > baseline). Paired-samples F-tests (or Wilcoxon signed-rank tests for non-normally distributed continuous variables) will be conducted to evaluate within-participant trajectories across the active treatment course for the entire cohort.

#### Analysis 2: Association between dietary intake and cardiometabolic outcomes

Multivariable linear regression models will be constructed to examine prospective associations between treatment-period dietary exposures and cardiometabolic endpoints. In the primary models (ANCOVA approach), the 6-month follow-up value of each endpoint will serve as the dependent variable, adjusted for its corresponding baseline value, age, baseline body mass index (BMI), tumor stage, cancer treatment regimen (chemotherapy alone vs. sequential HER2-directed targeted therapy), physical activity level at follow-up, and total energy intake (when modeling macronutrient percentage energy or nutrient densities). Secondary complementary models will evaluate change scores (Δ) as the dependent variable. Unstandardized regression coefficients (*B*) with 95% confidence intervals (CIs) and standardized coefficients (*β*) will be reported for all models.

#### Outcome hierarchy and interpretation

To protect against Type, I error inflation without compromising exploratory discovery, statistical inferences will adhere strictly to the prespecified outcome hierarchy defined in the study design. Formal confirmatory conclusions will be prioritized around the primary endpoint (visceral adiposity index; VAI). Key secondary endpoints (fat mass, HOMA-IR, and AST) and other secondary cardiometabolic parameters will be interpreted with appropriate caution as supportive and hypothesis-generating evidence, precluding formal adjustments for multiplicity.

#### Model diagnostics and missing data management

Multicollinearity among regression covariates will be evaluated using the variance inflation factor (VIF), with values < 5.0 considered acceptable. Key linear regression assumptions— including linearity, homoscedasticity, and normality of residuals—will be examined using standardized residual plots and normal Q–Q plots. Primary analyses will follow a complete-case approach, with the effective analytical sample size explicitly reported for each model. If missingness in outcomes or primary covariates exceeds 5% under the missing-at-random (MAR) assumption, multiple imputation by chained equations (MICE) with 20 imputed datasets will be implemented as a sensitivity analysis, pooling estimates according to Rubin’s rules to confirm the robustness of the complete-case results

## Discussion

The present prospective cohort study is designed to evaluate dietary intake patterns during the course of active cancer treatment in women with breast cancer and to investigate their associations with longitudinal cardiometabolic, biochemical, and psychological outcomes. By collecting repeated 24-hour dietary records across the early, middle, and concluding phases of treatment (up to the 6-month follow-up), this study aims to capture temporal variations in nutritional intake more accurately than previous studies that rely on a single dietary assessment. This comprehensive longitudinal approach is expected to provide a deeper understanding of patients’ habitual nutritional behaviors throughout treatment, ultimately helping to clarify how dietary exposure relates to treatment-induced cardiometabolic alterations and psychological well-being.

In addition, the study comprehensively evaluates multiple outcomes, spanning anthropometric indices, lipid and glycemic profiles, liver enzymes, and health-related quality of life. Evaluating these endpoints within a prespecified outcome hierarchy may help elucidate the complex interactions between diet, metabolic dysregulation, and psychological status in oncology care. Understanding these relationships will contribute to developing targeted nutritional interventions aimed at mitigating cardiometabolic toxicity and preserving quality of life among women undergoing breast cancer treatment.

This study possesses several notable strengths. Its prospective cohort design enables clear temporal assessment between dietary exposures during treatment and subsequent health outcomes at the 6-month follow-up. Furthermore, the protocol of collecting nine repeated 24-hour dietary records significantly mitigates the influence of day-to-day intake variability and captures true habitual consumption during cancer therapy. Additionally, the pre-specified outcome hierarchy designating the visceral adiposity index (VAI) as the primary endpoint— minimizes the risk of Type I error inflation while systematically exploring secondary cardiometabolic and psychological outcomes.

Several limitations warrant consideration. First, recruitment is restricted to selected oncology centers in Tehran, which may constrain the generalizability of the findings to breast cancer populations from diverse geographic, socioeconomic, and dietary backgrounds. Second, while the sample size (*n* = 100) was specifically powered for the primary outcome (VAI), statistical power may be modest for detecting subtle changes across certain secondary endpoints. Third, although dietary records were rigorously reviewed by trained nutritionists using food models and photographic atlases, self-reported dietary data remain susceptible to recall bias, underreporting, and misestimation of portion sizes. Fourth, completing nine dietary records during cytotoxic therapy imposes a notable participant burden; treatment-related side effects such as nausea, chronic fatigue, and cognitive fluctuations may lead to participant fatigue or missing entries. Finally, despite adjusting for key confounders—including age, baseline BMI, tumor stage, treatment regimen, and physical activity—the potential for residual confounding from unmeasured clinical factors, concomitant supportive medications, or genetic predispositions cannot be entirely ruled out.

## Data Availability

The datasets generated and analyzed during the current study are not publicly available due to patient privacy and ethical restrictions, but are available from the corresponding author on reasonable request.

## Acknowledgments

The authors used artificial intelligence–assisted tools to improve the clarity and readability of the manuscript. The authors reviewed, edited, and approved all content and assume full responsibility for the final version of the manuscript.

The Ethics committee/IRB of Shahid Beheshti University of Medical Sciences gave ethical approval for this work (Ethics code: IR.SBMU.CRC.REC.1402.021). Written informed consent will be obtained from all participants prior to their inclusion in the study.

## Authors’ Contributions

O.A. conceived and designed the study, developed the study protocol, and drafted the manuscript. M.E.A. and S.H.D. supervised the study and contributed to the development of the study design and critical revision of the manuscript. H.R.M. provided clinical and oncological expertise and contributed to patient identification and referral. B.N. provided statistical consultation and contributed to the methodological aspects of the study. All authors reviewed, revised, and approved the final version of the manuscript.

## Availability of Data and Materials

The datasets used and analyzed during the current study will be available from the corresponding author on reasonable request once the study is completed.

## References

1. Sung H, Ferlay J, Siegel RL, et al. Global cancer statistics 2020: GLOBOCAN estimates of incidence and mortality worldwide for 36 cancers in 185 countries. CA: a cancer journal for clinicians 2021;71(3):209–49.

2. Sun Y-S, Zhao Z, Yang Z-N, et al. Risk factors and preventions of breast cancer. International journal of biological sciences 2017;13(11):1387.

3. Russo S, Cinausero M, Gerratana L, et al. Factors affecting patient’s perception of anticancer treatments side-effects: an observational study. Expert opinion on drug safety 2014;13(2):139– 50.

4. Chala E, Manes C, Iliades H, et al. Insulin resistance, growth factors and cytokine levels in overweight women with breast cancer before and after chemotherapy. Hormones (Athens) 2006;5(2):137–46. doi: 10.14310/horm.2002.11177

5. Guinan EM, Connolly EM, Healy LA, et al. The development of the metabolic syndrome and insulin resistance after adjuvant treatment for breast cancer. Cancer Nurs 2014;37(5):355–62. doi: 10.1097/NCC.0b013e3182a40e6d

6. Arpino G, De Angelis C, Buono G, et al. Metabolic and anthropometric changes in early breast cancer patients receiving adjuvant therapy. Breast Cancer Res Treat 2015;154(1):127–32. doi: 10.1007/s10549-015-3586-x [published Online First: 20150930]

7. Alacacioglu A, Kebapcilar L, Gokgoz Z, et al. Leptin, insulin and body composition changes during adjuvant taxane based chemotherapy in patients with breast cancer, preliminary study. Indian J Cancer 2016;53(1):39–42. doi: 10.4103/0019-509x.180836

8. Bicakli DH, Varol U, Degirmenci M, et al. Adjuvant chemotherapy may contribute to an increased risk for metabolic syndrome in patients with breast cancer. J Oncol Pharm Pract 2016;22(1):46–53. doi: 10.1177/1078155214551315 [published Online First: 20140917]

9. Coskun T, Kosova F, Ari Z, et al. Effect of oncological treatment on serum adipocytokine levels in patients with stage II-III breast cancer. Mol Clin Oncol 2016;4(5):893–97. doi: 10.3892/mco.2016.815 [published Online First: 20160310]

10. Dieli-Conwright CM, Wong L, Waliany S, et al. An observational study to examine changes in metabolic syndrome components in patients with breast cancer receiving neoadjuvant or adjuvant chemotherapy. Cancer 2016;122(17):2646–53. doi: 10.1002/cncr.30104 [published Online First: 20160524]

11. Gepner Y, Shelef I, Schwarzfuchs D, et al. Effect of Distinct Lifestyle Interventions on Mobilization of Fat Storage Pools: CENTRAL Magnetic Resonance Imaging Randomized Controlled Trial. Circulation 2018;137(11):1143–57. doi: 10.1161/circulationaha.117.030501 [published Online First: 20171115]

12. Gomes SL, Bobby Z, Ganesan P, et al. Metabolic syndrome and its related biochemical derangements in breast cancer patients who received neoadjuvant chemotherapy: A study from a tertiary care oncology centre from Puducherry, South India. Diabetes Metab Syndr 2021;15(3):975–80. doi: 10.1016/j.dsx.2021.04.022 [published Online First: 20210427]

13. A A, Sude NS, B AR, et al. Prospective Evaluation of Response Outcomes of Neoadjuvant Chemotherapy in Locally Advanced Breast Cancer. Cureus 2022;14(2):e21831. doi: 10.7759/cureus.21831 [published Online First: 20220202]

14. Kim HY, Ju S, Jung YJ, et al. Effects of Chemotherapy (Anthracyclin, Cyclophosphamide following Docetaxel Regimen) on Sleep, Anxiety, Depression, and Quality of Life in Patients with Breast Cancer. Oncology 2025;103(11):974–84. doi: 10.1159/000543730 [published Online First: 20250130]

15. Arends J, Bachmann P, Baracos V, et al. ESPEN guidelines on nutrition in cancer patients. Clin Nutr 2017;36(1):11–48. doi: 10.1016/j.clnu.2016.07.015 [published Online First: 20160806]

16. Custódio ID, Marinho Eda C, Gontijo CA, et al. Impact of Chemotherapy on Diet and Nutritional Status of Women with Breast Cancer: A Prospective Study. PLoS One 2016;11(6):e0157113. doi: 10.1371/journal.pone.0157113 [published Online First: 20160616]

17. de Kruif AJ, Westerman MJ, Winkels RM, et al. Exploring changes in dietary intake, physical activity and body weight during chemotherapy in women with breast cancer: A Mixed-Methods Study. J Hum Nutr Diet 2021;34(3):550–61. doi: 10.1111/jhn.12843 [published Online First: 20210107]

18. Gezalan S, Ateş S. The impact of taste and smell alterations on nutritional status and quality of life in individuals undergoing chemotherapy. Support Care Cancer 2025;33(11):986. doi: 10.1007/s00520-025-09960-2 [published Online First: 20251028]

19. James S, Oppermann A, Schotz KM, et al. Nutritional Counseling During Chemotherapy Treatment: A Systematic Review of Feasibility, Safety, and Efficacy. Current Oncology 2025; 32(1).

20. Vella R, Pizzocaro E, Bannone E, et al. Nutritional Intervention for the Elderly during Chemotherapy: A Systematic Review. Cancers 2024; 16(16).

21. Schwingshackl L, Hoffmann G. Adherence to Mediterranean diet and risk of cancer: an updated systematic review and meta-analysis of observational studies. Cancer Med 2015;4(12):1933–47. doi: 10.1002/cam4.539 [published Online First: 20151016]

22. Wayne SJ, Baumgartner K, Baumgartner RN, et al. Diet quality is directly associated with quality of life in breast cancer survivors. Breast Cancer Res Treat 2006;96(3):227–32. doi: 10.1007/s10549-005-9018-6

23. Toloi JM, Laranja ACG, Toledo DO, et al. The Effects of Higher Protein Intake on Muscle Mass and Clinical Outcomes in Critically Ill Cancer Patients: A Prespecified Per-Protocol Analysis. Nutrients 2025;17(17) doi: 10.3390/nu17172742 [published Online First: 20250824]

24. Wang J, Wu SG. Breast Cancer: An Overview of Current Therapeutic Strategies, Challenge, and Perspectives. Breast Cancer (Dove Med Press) 2023;15:721–30. doi: 10.2147/bctt.S432526 [published Online First: 20231020]

25. Freihat O, Sipos D, Kovacs A. Global burden and projections of breast cancer incidence and mortality to 2050: a comprehensive analysis of GLOBOCAN data. Front Public Health 2025;13:1622954. doi: 10.3389/fpubh.2025.1622954 [published Online First: 20251030]

26. Nurgali K, Jagoe RT, Abalo R. Editorial: Adverse Effects of Cancer Chemotherapy: Anything New to Improve Tolerance and Reduce Sequelae? Front Pharmacol 2018;9:245. doi: 10.3389/fphar.2018.00245 [published Online First: 20180322]

27. de Vries YC, van den Berg M, de Vries JHM, et al. Differences in dietary intake during chemotherapy in breast cancer patients compared to women without cancer. Support Care Cancer 2017;25(8):2581–91. doi: 10.1007/s00520-017-3668-x [published Online First: 20170316]

